# SARS-CoV-2 transmission from the healthcare setting into the home: a prospective longitudinal cohort study

**DOI:** 10.1101/2021.02.01.21250950

**Authors:** Simon Craxford, Jessica Nightingale, Ben Marson, Amrita Vijay, Alan Norrish, Adeel Ikram, Lola M L Cusin, Patrick Tighe, Guruprasad P Aithal, Stuart Astbury, Jonathan K. Ball, Jayne Newham, Richard A. Urbanowicz, Anthony Kelly, Waheed Ashraf, Alexander W. Tarr, Ana M Valdes, Benjamin J Ollivere

## Abstract

**Objective:** To assess the incidence of symptomatic and asymptomatic SARS-CoV-2 seropositivity in healthcare workers and subsequent transmission to their close contacts within their household. To assess changes in immunoglobulin (Ig) and neutralising antibodies (nAbs) in exposed participants.

**Setting:** Two acute National Health Service (NHS) hospitals within the East Midlands region of England.

**Background:** The UK has been one of the most severely affected countries during the COVID-19 pandemic. Transmission from healthcare workers to the wider community is a potential major vector for spread of SARS-CoV-2 which is not well described in the current literature.

**Methods:** Healthcare workers (HCW) were recruited from two Hospitals within the East Midlands of England and underwent serial blood sampling for anti-SARS-CoV-2 antibodies (both nucleocapsid and spike protein for IgG, IgM and IgA) between 20 April and 30 July 2020, with the presence of neutralising antibodies (nAbs) assessed for positive participants. Cohabitees of the volunteers were invited to attend testing in July -August 2020 and underwent identical serological testing as the HCWs.

**Results:** 633 healthcare professionals were recruited. 178 household contacts of 137 professionals volunteered for the study. 18% of healthcare professionals (115 out of 633) tested as seropositive during the study period, compared to an estimated seroprevalence of 7% within the general population. The rate of symptomatic COVID-19 was 27.5% compared to an asymptomatic rate of 15.1%. Rates of positivity declined across the study period for all immunoglobulins (overall positivity from 16.7% to 6.9%).

7.2% of the cohabitees tested as seropositive. 58 cohabitees lived with a serologically positive HCW; this group had a seropositive rate of 15.5%, compared to 2.5% of cohabitees without a seropositive HCW, a six-fold increase in risk (Odds ratio 7.16 95% CI 1.86 to 27.59), p = 0.0025). Given the observed decay rates and data from Public Health England, we estimate that the proportion of seropositive cohabitees living with a seropositive HCW at the height of the first wave could have been as high as 44%.

110 out of 115 (95.7%) HCWs and 12 out of 13 (92.3%) cohabitees who tested positive developed detectable nAbs. 56.5% (65 out of 115) of SARS-CoV-2 positive HCWs developed a neutralising titre with an IC_50_≥1/300; no cohabitee achieved this level..

**Conclusions:** Transmission of SARS-CoV-2 between healthcare professionals and their home contacts appears to be a significant factor of viral transmission, but, even accounting for the decline in seropositivity over time, less than 44% of adult cohabitees of seropositive healthcare workers became seropositive. Routine screening and priority vaccination of both healthcare professionals and their close contacts should be implemented to reduce viral transmission from hospitals to the community.

**SUMMARY BOXES:** *Section 1: What is already known on this topic:*

- Healthcare workers (HCWs) have increased rates of SARS-CoV-2 infection compared with the general population due, at least in part, to high levels of occupational exposure.
- IgA, IgM and IgG are detectable for most patients after 11 days post SARS-CoV-2 infection but all decline in the weeks following SAR-CoV-2 exposure.
- Rates of transmission to healthcare workers, and therefore subsequent transmission to their close contacts, may be reduced with effective PPE.

*Section 2: What this study adds:*

- The amount of neutralising antibodies formed may be dependent on IgG response as it is much lower among seropositive cohabitees than seropositive healthcare workers.
- NHS Healthcare workers had a far greater seroprevalence of SARS-CoV-2 infection compared to the general population.
- Cohabitees of positive healthcare workers have a 6-fold increased risk of developing serological evidence of SARS-CoV-2 infection compared to the general population.
- Despite this increased risk, transmission at home is less than 50% even from highly exposed healthcare workers, but remains an important potential vector of transmission from hospitals to the wider community.

**Research into context:** *Evidence before this study:* We searched PubMed for articles published between January 1 2020 and January 27, 2021 with the terms “Covid-19”, “healthcare workers”, and “transmission” “home {NOT nursing} or household”. We did not restrict our search by language or type of publication. We identified 38 studies of which only one assessed the prevalence among HCW households using Canadian national databases. Our PubMed search yielded only one serological study within the German Healthcare system, which suggested very low transmission from healthcare workers to their close cohabitees.

*Added value of this study:* To our knowledge, this is the largest longitudinal serological cohort study assessing transmission of SARS-CoV-2 infection from the UK healthcare environment to the home (n = 633 healthcare workers, 178 cohabitees). Our findings showed that serological evidence within the HCW was high with 18% of healthcare professionals (115 out of 633) tested as seropositive during the study period, compared to an estimated seroprevalence of 7% within the general population. A cohabitee of a seropositive HCW had a six-fold increase of being seropositive themselves compared to a baseline rate of 2.5%. Despite this increased risk, transmission at home is less than 50% even from highly exposed healthcare workers, but remains an important potential vector of transmission from hospitals to the wider community. Rates of positivity declined across the study period for all immunoglobulins (overall positivity from 16.7% to 6.9%). Given the observed decay rates and data from Public Health England, we estimate that the proportion of seropositive cohabitees living with a seropositive HCW at the height of the first wave could have been as high as 44%.

*Implications of all available evidence:* Understanding the transmission during the first wave from the healthcare setting into the home and the extent of such transmissions is essential to understand containment strategies of novel SARS-CoV-2 variants or to understand viral transmission of future respiratory viruses. NHS workers appeared to be at an increased risk of contracting of SARS-CoV-2 infection compared to the HCWs of other nations; we hypothesise that this may be related to a scarcity of appropriate personal protective equipment during the initial wave of SARS-CoV-2. Healthcare workers (HCWs) have increased rates of SARS-CoV-2 infection compared with the general population. An infected HCW, whether symptomatic or not, appears to be a significant bridge for transmission of SARS-CoV-2 to their close home contacts.

## Background

The United Kingdom (UK) has been severely affected by both the first and current second wave of severe acute respiratory syndrome coronavirus 2 (SARS-CoV-2), which causes COVID-19^1^.

The transmission of SARS-CoV-2 is mainly considered to occur via person-to-person close contact by droplet infection ^2^. Front line healthcare workers (HCWs) responding to the current pandemic have many more close contacts with potential COVID-19 patients and so are at an elevated risk of contracting COVID-19 compared to the general population ^3^. Published studies suggest that front-line healthcare workers could account for up to 20% of all COVID-19 diagnoses ^4 5^. A lack of adequate personal protection equipment (PPE) and delays in setting up an effective contact tracing system during the first wave of the pandemic may also contribute to this increased risk ^6,7^.

Current UK guidelines advise household contacts to isolate within the same home as the index case for 10 days following a positive test^8^. Despite guidance advising household members to socially distance, cohabitees of the affected household are likely to interact repeatedly, including in the use of shared facilities such as bathrooms or kitchens, and therefore be exposed to potential SARS-CoV-2 transmission^9^. Household cohabitees make up the majority of close contacts for infected individuals and may account for up to 70% of SARS-CoV-2 transmissions when widespread community measures, such as local lockdowns, are in place ^10^. As healthcare providers are designed as key workers who attend their job despite COVID-19 restrictions, asymptomatic transmission within this group may also undermine the effects of nationally instituted “lock-downs”. Hospitals, healthcare workers and their cohabitees may therefore provide an ongoing reservoir of SARS-CoV-2 infection ^3 11^.

Studies assessing transmission of SARS-CoV-2 have so far focused on symptom trackers and single serology samples ^12^. However, these studies may miss asymptomatic infections. Few prospective longitudinal serological studies of healthcare workers have been performed to date.

The primary aim of this study was to assess rates of both symptomatic and asymptomatic seropositivity within a patient-facing healthcare worker (HCW) population and subsequent transmission from HCW to their household cohabitees. The longitudinal design of the study also allowed for the characterisation of Ig response and formation of neutralising antibodies during the first wave of SARS-CoV-2.

## Methods

### Recruitment

The PANdemic Tracking of Healthcare workers (Panther) prospective cohort began in April 2020. Between April 24^th^ and July 10^th^ 2020, patient-facing healthcare professionals were recruited from the Queen’s Medical Centre and City Hospital located in Nottingham, UK and formed the “Healthcare workers Cohort”. Participants included asymptomatic healthcare workers and support staff working at these hospitals who were required to complete a health questionnaire (attached as a Supplementary file) and provide blood samples at 9 study visits (baseline, weeks 1, 2, 3, 4, 6, 8, 10 and at month 4 and month 6 following recruitment). All participants provided written informed consent prior to completing the questionnaire and providing blood samples. Blood samples at each visit was collected using BD Serum Separator Tubes (SST-II) for serum. PAX gene Vacutainer tubes for RNA and EDTA tubes for DNA were collected at baseline. Bloods collected for serum were processed within 30 minutes of collection and stored at −80°C until further analysis. Bloods for RNA and DNA extractions were collected and stored immediately at − 20°C for the first 24 hours and then transferred to −80°C. Our study website (https://pantherstudy.org.uk/) was primarily developed as the gateway to increase participation rate by providing background information on the aims and objectives of the study and how participants could enrol into the study.

In September 2020, study participants were invited to bring their household contacts (aged over 18) for a single blood sample. These participants were invited at the earliest opportunity when they could safely attend the hospital for blood testing. These recruits formed the “Household contacts” cohort.

The PANTHER protocol is available online (www.pantherstudy.org.uk). Samples were collected and stored under a Human Tissue Authority licence in Nottingham Tissue Bank (Licence number: 11035). The study protocol was approved by North West - Greater Manchester South Research Ethics Committee (reference 20/NW/0395).

### Serological processing

#### Anti-S1 and anti-nucleocapsid ELISA

Serum samples were serially diluted in 3% skimmed milk powder in PBS containing 0.05% Tween 20 and 0.05% sodium azide. All assays were performed on Biotek Precision liquid handling robots in a class II microbiological safety cabinet. For endpoint dilution ELISAs, sera were progressively 4-fold diluted from 1:150 to 1;38,400. ELISA was performed by coating 384 well Maxisorp (NUNC) assay plates with either 20 µL per well of 0.5 µg.mL-1 of Wuhan strain SARS-CoV-2 spike protein S1 subunit (His tagged, HEK293 expressed; Sino Biological) or SARS-CoV-2 nucleocapsid (His Tagged, baculovirus expressed; Sino Biological) in carbonate-bicarbonate buffer (CBC; Merck), or human IgG at 1 µg.mL-1 in CBC buffer as controls. Plates were sealed with foil film and incubated overnight at 4 °C. Plates were then washed with PBS with 0.05% Tween 20 (PBS-T) 3 times using a ThermoFisher Wellwash Versa plate washing robot. Wells were immediately filled with 100 µL of 3% skimmed milk powder (w/v) in PBS and 0.05% sodium azide (PBS-MA) and blocked overnight at 4 °C. Assay plates were then washed 3 times and 20 µL of pre-diluted serum sample (including SARS-CoV-2 antibody-positive and negative serum controls) added in duplicate wells. After one hour at 21 °C, the plate was washed 3 times in PBS-T, followed by addition of 20 µL of gamma chain-specific anti-human IgG-HRP conjugate (Sigma A0170-1ML) at 1:30,000 dilution, incubating for one hour at 21 °C. Following a final three washes with PBST, 40 µL One-step UltraTMB substrate solution (ThermoScientific) was added to each well. After incubating for 20 minutes at room temperature, 40 µL of 2N H2SO4 was added to each well and Absorbance was measured at 450nm using a GlowMax Explorer microplate reader (Promega).

#### SARS-CoV-2 pseudotype neutralization assay

Participants found to be seropositive for SARS-CoV-2 were assessed for the presence of neutralising antibodies. The latest positive time point was utilised for each participant for neutralising antibody assays to standardise results in evolving immunity. Human Embryonic Kidney 293T and VeroE6 cell lines were purchased from ECACC. Both cell lines were grown in Dulbecco’s modified eagle medium (DMEM; Invitrogen), supplemented with 10% fetal bovine serum (Invitrogen), and 0.1mM nonessential amino acids (GIBCO) at 37°C and 5% CO2. Cell lines were tested to verify the absence of mycoplasma. No assays were performed to verify the identity of the cells in culture following receipt of the cells.

Pseudotypes were produced as previously described ^13^. Briefly, 1.5 × 106 HEK293T cells were seeded overnight in a 10 cm diameter Primaria-coated dish (Corning). Transfections were performed by mixing 2 µg of murine leukemia virus (MLV) Gag-Pol packaging vector (phCMV-5349), 2 µg of luciferase encoding reporter plasmid (pTG126) and 2 µg Wuhan strain SARS-CoV-2 spike plasmid with 24 µL polyethylenimine (Polysciences) in Optimem (Gibco). Mixtures were added to cells for 6 hours, after which they were replaced with complete DMEM. A no-envelope control (nude pseudotype) was used as a negative control in all experiments. Supernatants containing SARS-CoV-2 pseudotype viruses (pv) were harvested 72 h post-transfection and filtered through 0.45 μm membranes. For infectivity and neutralization testing of SARS-CoV-2 pv, 2 × 10^4^ VeroE6 cells/well were plated in white 96-well tissue culture plates (Corning) and incubated overnight at 37 °C. The following day, SARS-CoV-2 pv-containing supernatants were incubated with 1/300 dilution of sera (pre-treated with 1% Triton X-100/heat-inactivated for 1 hr at RT), before being added to VeroE6 cells for 4 h. Infection inocula were then discarded and 200 µL DMEM added to the cells. After 72 h, media was discarded, cells lysed with cell lysis buffer (Promega) and relative luciferase activity (in RLU) measured using a FLUOstar Omega plate reader (BMG Labtech). Each sample was tested in triplicate.

### Statistical analysis

Univariable and multivariable binary logistic regression was performed to identify co-variables contributing to a positive sample. The Index of Multiple Deprivation (IMD) decile for their geographical area was assigned using the participant’s post code. Other variables for adjustment include age, sex, ethnicity, symptoms of COVID-19 and job role. Participants were classified as seropositive if they had a positive titre to either the NC or the S1 protein at any of the time-points sampled. Participants were classified as having reported symptoms if at any time point during the period of assessment if they complained of a symptom attributable to suspected viral infection (headaches, fever, cough, loss of sense of smell or taste, fatigue, difficulty breathing) or they had to self-isolate with suspected COVID-19. Estimates of seroprevalence for the general population were obtained using data from Public Health England^14^]. Seropositive cohabitees with a negative HCW were used as a “control” to estimate the likely decay in positivity and the likely “true” rate of cohabitee infection earlier during the pandemic.

Neutralising antibodies were stratified based on achieving a neutralising percentage of 50% or greater at 1/300 dilution. The relationship with peak IgG, IgA or IgM and neutralisation percentage was assessed using multiple linear regression, and predictors influencing neutralisation percentage was assessed using binary multivariable logistic regression / multiple linear regression. Lastly, neutralising percentages were correlated between HCWs and cohabitees using Spearman’s rho.

Statistical analysis was performed using IBM SPSS Version 24. A p value of <0.05 was deemed statistically significant. Figures were produced using Graphpad Prism version 8.

## Results

### Sample

633 healthcare professionals (HCWs) were recruited across the two acute hospitals. 178 household contacts from 137 HCWs volunteered for the study. The recruitment is shown in Figure 1.

**Figure 1:**
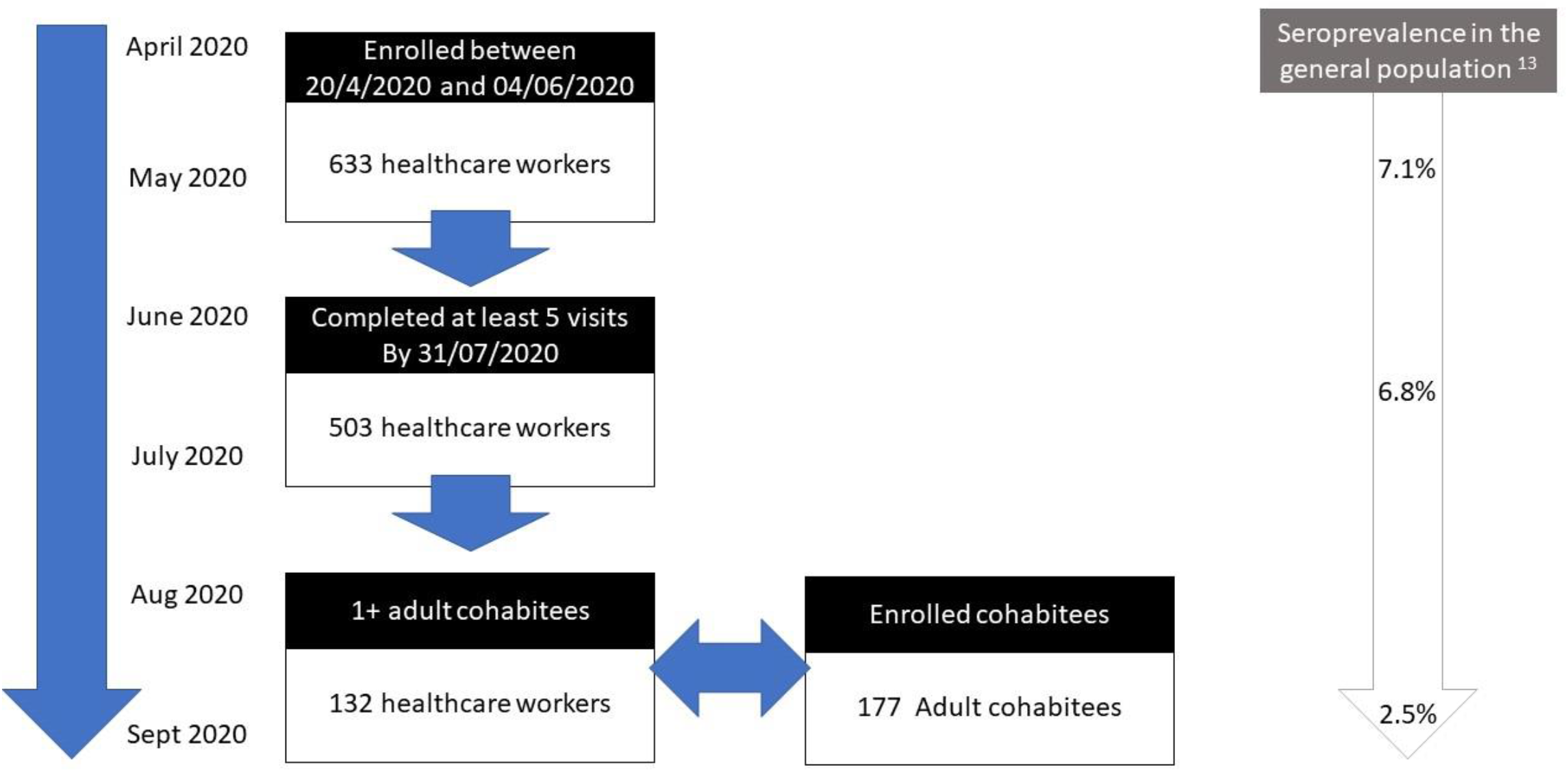
Consort diagram for the Panther study. Seroprevalence for the Midlands is estimated from Public Health England data ^14^ and from cohabitee data.

Figure 1: Study recruitment, seropositivity and estimated seropositivity of population of the Midlands ^14^. (see end of manuscript).

22 HCWs (3.48%) reported testing positive on a nasal swab COVID PCR either during the study period or immediately before. HCWs inviting their contacts to attend the study had a greater rate of seropositivity, with a rate of 37%. The cohabitee contacts had an overall 7.2% seropositivity. Of the 58 cohabitees of seropositive HCW had a seropositive rate of 15.5%, compared to 2.5% of cohabitees without a seropositive HCW (p = 0.0025, Fishers Exact). While 12 out of 13 samples demonstrated detectable neutralising antibodies, no cohabitee displayed neutralising antibodies of greater than 50%.

No participant required hospital treatment for COVID-19 during the study, however the spouse of one participant died of COVID-19 before the start of the study. The demographic characteristics of the cohorts are summarized in table 1.

**Table 1:** Demographics and proportion of ‘anti-SARS-CoV-2 nucleocapsid positive participants for healthcare workers and cohabitees.

|  |  | N anti-NC<br>negative | N anti-NC<br>positive | % anti-NC<br>positive |
| --- | --- | --- | --- | --- |
| **Healthcare workers** |  |  |  |  |
| **Participants** | Total | 518 | 115 | 18.2% |
| **Age** | Mean years<br>(SD) | 43.1<br>(11.6) | 43.9<br>(11.6) |  |
| **Gender** | M | 115 | 22 | 16.1% |
|  | F | 401 | 91 | 18.7% |
|  | Other/Not specified | 2 | 0 | 0.0% |
| **Symptomatic** | yes | 108 | 42 | 28.0% |
|  | no | 410 | 73 | 15.1% |
| **Ethnicity\*** | South Asian | 40 | 10 | 20.0% |
|  | East Asian | 6 | 2 | 25.0% |
|  | Black | 10 | 7 | 41.2% |
|  | Mixed | 24 | 9 | 27.3% |
|  | Not stated | 1 | 0 | 0.0% |
|  | White | 439 | 87 | 16.5% |
| **IMD decile** | 1-3 | 96 | 29 | 23.2% |
|  | 4-7 | 177 | 38 | 17.7% |
|  | 8-10 | 130 | 29 | 18.2% |
|  | Unspecified | 113 | 19 | 14.4% |
| **Hospital role** | ITU role | 36 | 1 | 2.7% |
|  | Frontline not ITU role | 484 | 114 | 19.1% |
| **Medical co-morbidity** | Smoker | 116 | 22 | 15.9% |
|  | Diabetes | 9 | 0 | 0.0% |
|  | Respiratory disease | 62 | 18 | 22.5% |
|  | Cardiovascular disease | 2 | 9 | 18.2% |
|  | Hypertension | 8 | 33 | 19.5% |
| **Cohabitees** |  |  |  |  |
| **Participants** | Total | 165 | 13 | 7.30% |
| **Linked HCW Status** | Positive HCW | 49 | 9 | 15.5% |
|  | Negative HCW | 117 | 3 | 2.6% |
| **Age** | Mean years (SD) | 38.31<br>(15.72) | 41.41<br>(14.68) |  |
| **Gender** | Male | 46 | 9 | 16.4% |
|  | Female | 119 | 3 | 2.5% |
| **BMI** | Mean (SD) | 26.0 (5.2) | 26.3 (4.7) |  |
|  | Did not state | 0 | 1 | 100% |
| **Symptoms** | Symptomatic | 24 | 3 | 11.1% |
|  | Asymptomatic | 141 | 9 | 6.0% |
| **Ethnicity\*** | South Asian | 5 | 1 | 16.7% |
|  | East Asian | 5 | 1 | 16.7% |
|  | Black | 6 | 0 | 0 |
|  | Mixed | 2 | 0 | 0 |
|  | Other | 0 | 1 | 100% |
|  | White | 147 | 10 | 6.4% |
| **Medical co-morbidity** | Smoker | 7 | 0 | 0 |
|  | Diabetes | 5 | 0 | 0 |
|  | Respiratory disease | 22 | 2 | 8.3% |
|  | Cardiovascular disease | 4 | 1 | 20% |
|  | Hypertension | 9 | 1 | 10% |

\*ethnicity self selected by participant from Office of National Statistics categories

**Table 2:** Unadjusted and adjusted Odds Ratios (OR) for risk factors for developing antibodies to COVID-19.

| Variable | Unad OR | 95% CI | P | Ad. OR | 95% CI | P |
| --- | --- | --- | --- | --- | --- | --- |
| Age | 1.005 | 0.99 to 1.02 | 0.57 | 1.01 | 0.989 to 1.026 | 0.43 |
| Symptomatic | 1.98 | 1.28 to 3.07 | **0.002** | 2.10 | 1.33 to 3.29 | **0.001** |
| ITU role | 0.38 | 0.11 to 1.25 | 0.112 | 0.33 | 0.099 to 1.121 | 0.076 |
| Male Gender | 0.76 | 0.46 to 1.30 | 0.334 | 0.80 | 0.466 to 1.36 | 0.404 |
| Smoker | 0.88 | 0.53 to 1.45 | 0.609 | 0.96 | 0.57 to 1.62 | 0.882 |
| Diabetes | 0.56 | 0.069 to 4.48 | 0.580 | 0.53 | 0.062 to 4.55 | 0.563 |
| Respiratory disease | 1.31 | 0.69 to 2.51 | 0.417 | 1.35 | 0.692 to 2.62 | 0.380 |
| Cardiovascular disease | 0.99 | 0.21 to 4.66 | 0.99 | 0.93 | 0.191 to 4.54 | 0.930 |
| Hypertension | 1.09 | 0.49 to 2.43 | 0.83 | 0.99 | 0.423 to 2.31 | 0.979 |
| Ethnicity |  |  |  |  |  |  |
| Black | 2.60 | 0.94 to 7.20 | 0.067 | 2.98 | 1.01 to 8.74 | **0.047** |
| East Asian | 1.30 | 0.51 to 3.29 | 0.582 | 1.46 | 0.56 to 3.78 | 0.446 |
| Mixed | 1.19 | 0.39 to 3.64 | 0.760 | 1.34 | 0.54 to 3.31 | 0.519 |
| South Asian | 1.28 | 0.54 to 3.04 | 0.573 | 1.36 | 0.54 to 3.31 | 0.532 |
| White | 1 |  |  | 1 |  |  |

### Serology

Overall, 18% of healthcare professionals (115 out of 633) tested as seropositive during the study period. 149 participants reported symptoms consistent with a viral infection; 41 had serological evidence of SARS-CoV-2 infected (27.5%). The rate of SARS-CoV-2 for HCWs without symptoms was 15.1% (73 out of 484). There was considerable variability within the exact immunoglobulin response generated by the participants after exposure to SARS-CoV-2. While the total number of people who had serological evidence of SARS-CoV-2 increased during the study period, rates of IgG, IgM and IgA positivity all showed significant decay, as shown in Figure 2.

**Figure 2:**
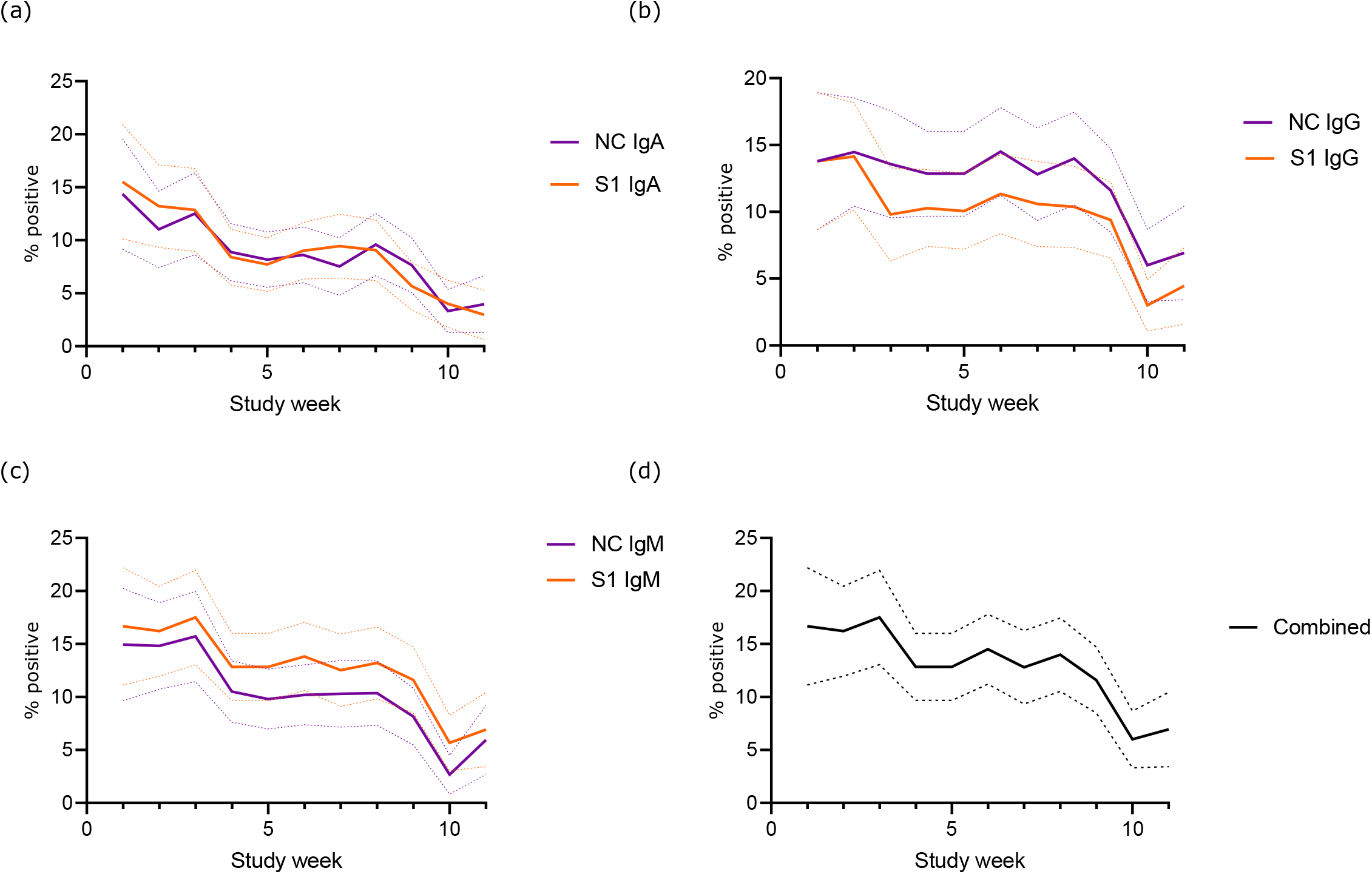
Proportion of participants testing positive to (a) IgA, (b) IgG and (c) IgM and d) Overall. Nuclear capsid rates are in purple and spike protein are in orange.

### Neutralising antibodies

110 out of 114 (96.5%) HCWs who had tested positive for any immunoglobulin had detectable neutralising antibodies at least two months following seropositivity. With our threshold of 50% neutralisation at a 1:300 triton dilution 57% (65 out of 114) of SARS-CoV-2 positive participants developed an effective neutralising antibody response.

Univariate analysis suggested that a higher peak of IgM Spike protein was associated with an increased chance of a sustained neutralising percentage of greater than 50%; this was not significant after multivariable adjustment (see figure 3).

**Figure 3:**
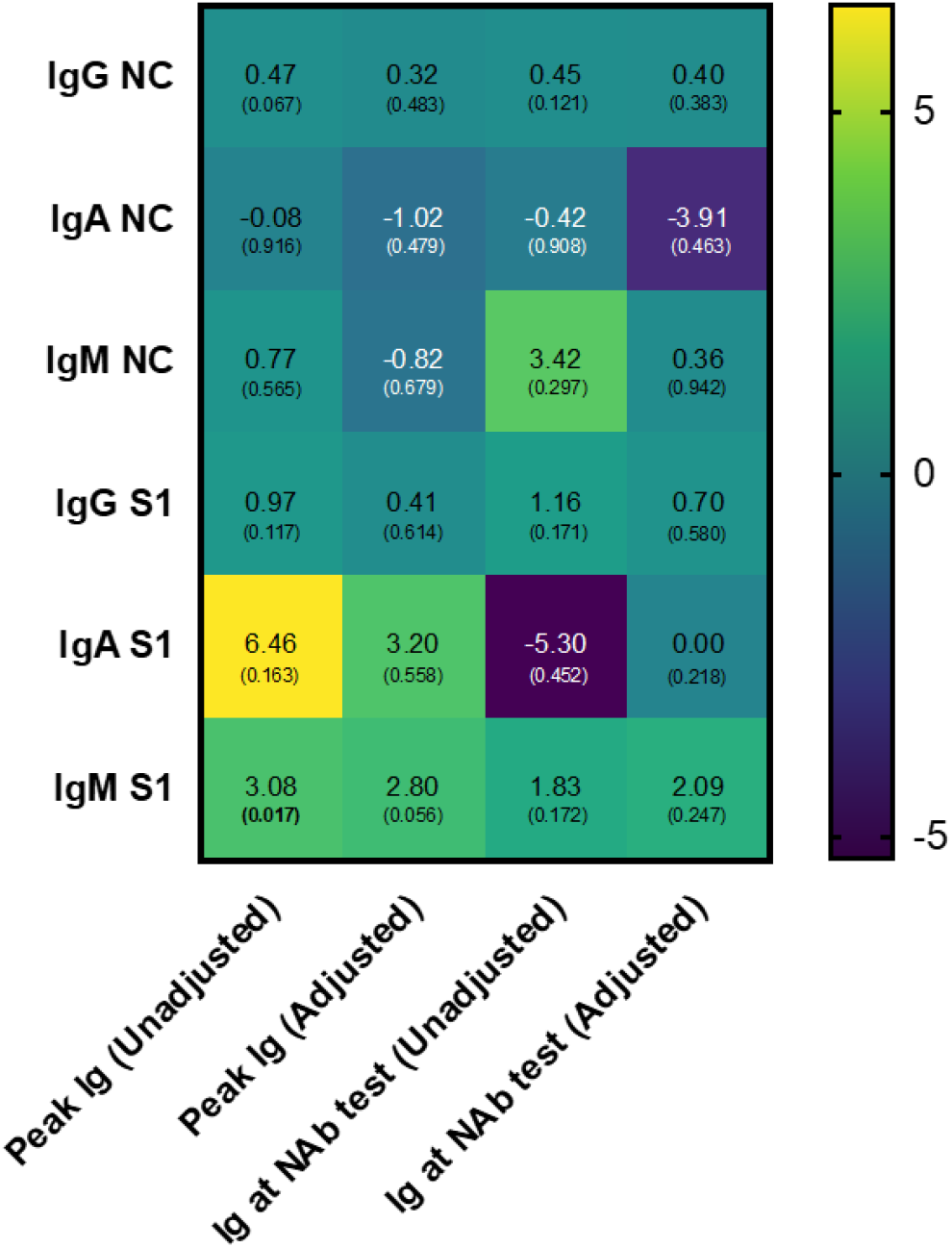
Heat map demonstrating unadjusted and adjusted Odds Ratios for immunoglobulin levels potentially influencing neutralising antibody formation.

Multiple linear regression was carried out to investigate whether peak spike protein Ig and ethnicity could significantly predict levels of neutralising antibodies. The results of the regression indicated that the model explained only 22.7% of variance and that the model was a significant predictor of neutralising antibody status, F =3.22, p = 0.021. Only peak IgG and white ethnicity contributed significantly to the model (see figure 4).

**Figure 4:**
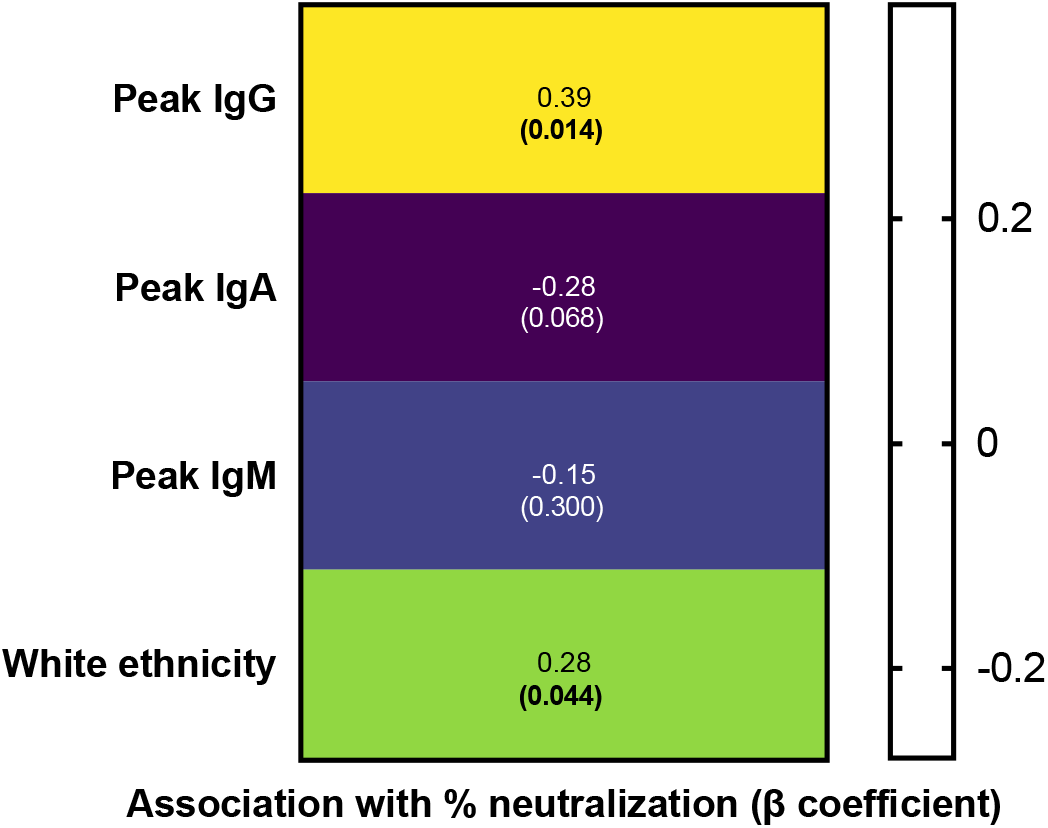
Heat map demonstrating predictors of neutralising antibody response

While regression modelling suggested a potential link between peak IgG and neutralising antibodies, several volunteers who had a high and sustained IgG response did not go on mount a neutralisation response of 50% or greater at 1/300 dilution (see figure 5).

**Figure 5:**
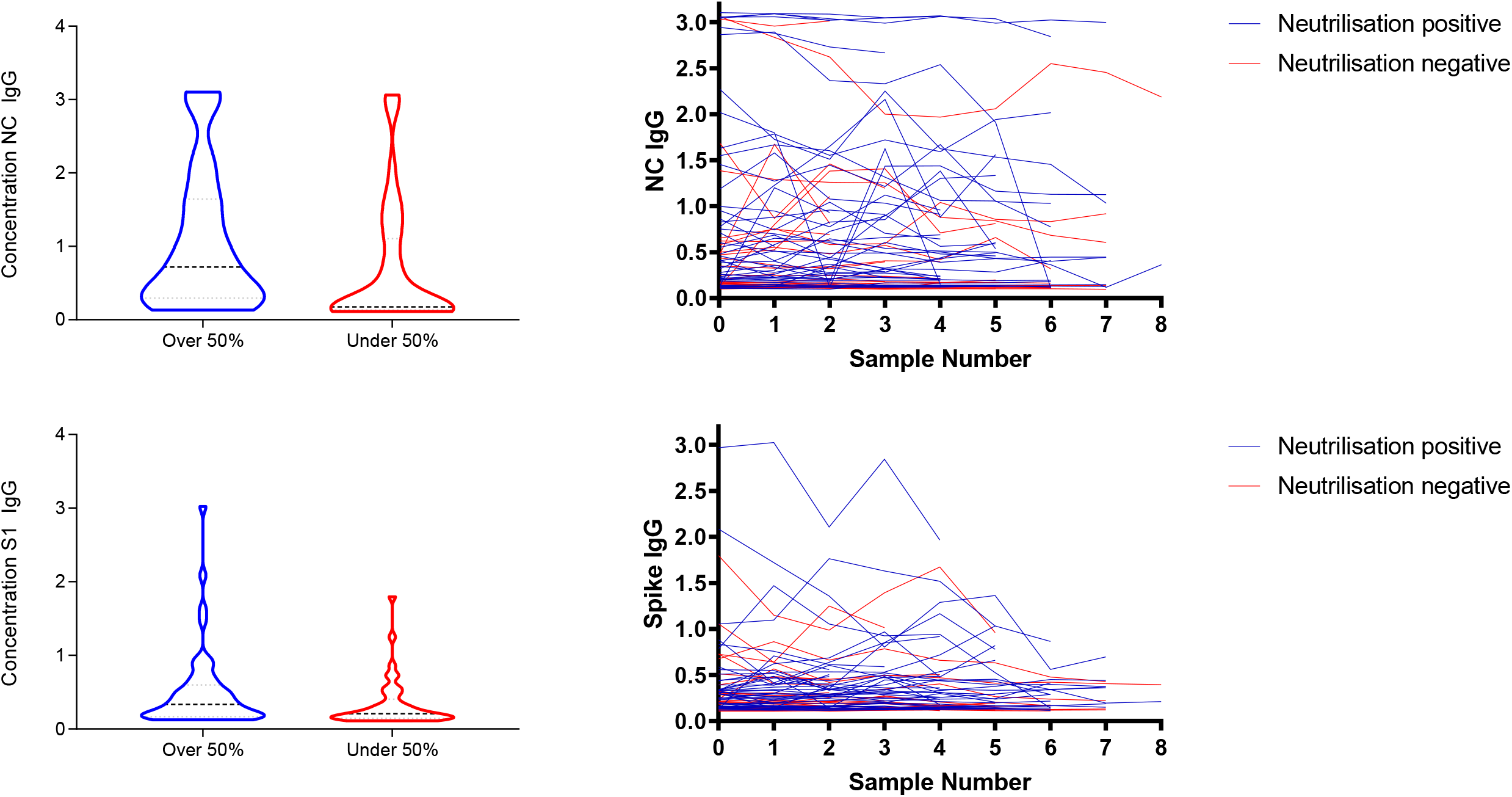
Mean IgG NC and S1 and serial values in relation to developing neutralising antibodies. Blue represents participants who achieved 50% or greater neutralisation while red represents those who did not.

178 household contacts from 137 HCWs volunteered for the study. HCWs inviting their contacts to attend the study had a greater rate of seropositivity, with a rate of 37%. The cohabitee contacts had an overall 7.2% seropositivity. Of the 58 cohabitees of seropositive HCW had a seropositive rate of 16%, compared to 2.5% of cohabitees without a seropositive HCW (p = 0.0025, Fishers Exact). This represents a six-fold increase in risk (unadjusted Odds ratio 7.16 95% CI 1.86 to 27.59, p = 0.0025, adjusted OR for ethnicity 6.86, CI 1.74 to 27.02).

While 12 out of 13 samples demonstrated detectable neutralising antibodies, no cohabitee displayed neutralising antibodies of greater than 50% (see figure 6). There was no strong correlation in neutralising antibody percentage between paired HCWs and cohabitees (Spearman’s rho correlation coefficient 0.311, p=0.301).

**Figure 6:**
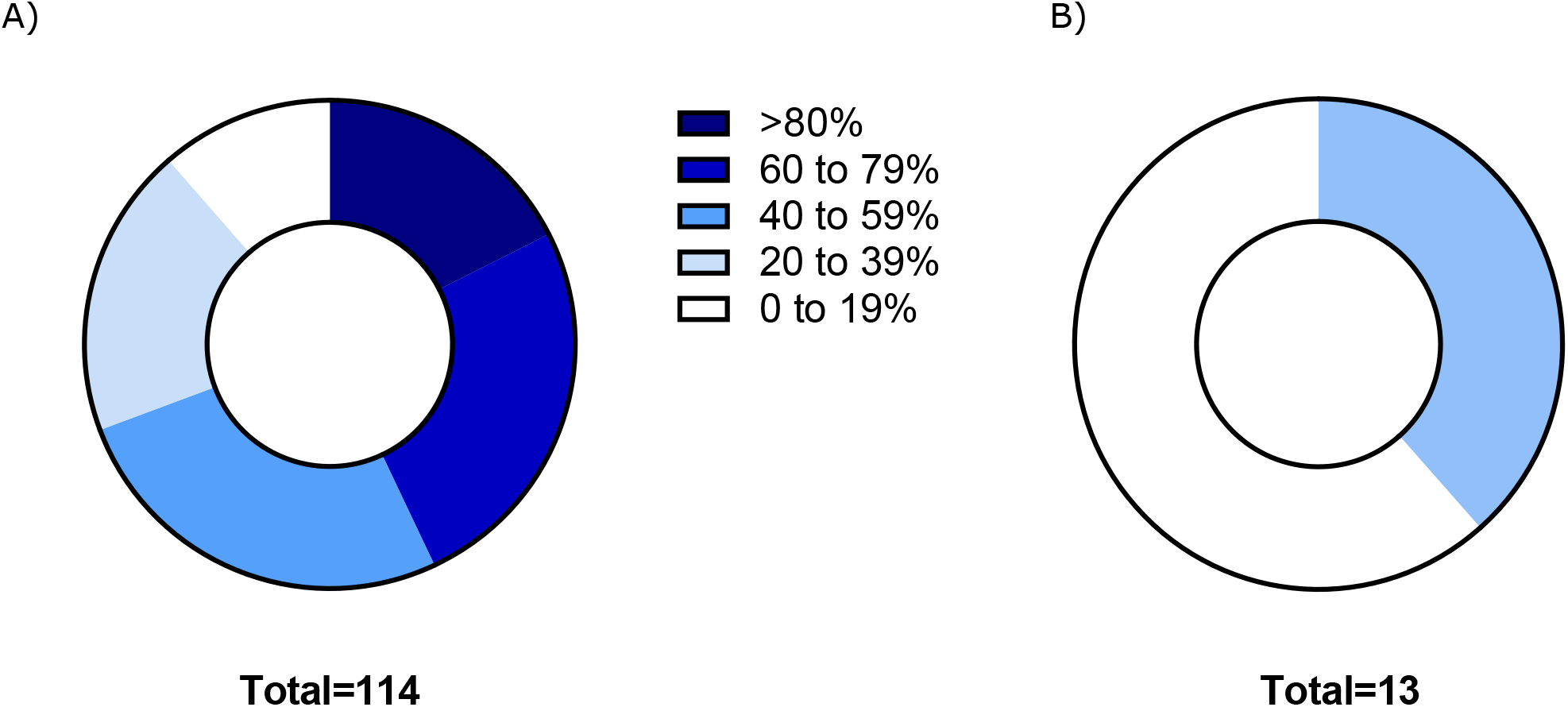
Potency of neutralising antibodies produced in A) Healthcare workers and B) cohabitees

### Antibody response to SARS-CoV-2

Univariable and multivariable analysis of risk factors for HCWs developing a serological response to SARS-CoV-2 in table 4. Only black ethnicity and the presence of symptoms were associated with an increased risk of SARS-CoV-2 seropositivity.

## Discussion

### Healthcare worker and cohabitee SARS-CoV-2 infection

Health care workers are at an elevated risk of infection as they work on the frontline of the response to the current pandemic. This study found that 18% of healthcare workers had antibody evidence of SARS-CoV-2 infection. This is higher than the estimated seropositive rate of the overall Midlands population during the same time period, which was between 6.5 and 7.2% throughout our study period^15^. Currently, few longitudinal serological studies of HCWs have published. Behrens et al and Herzberg et al found a lower serological rate of infection within German HCWs of between 1.8% and 4% respectively, which was attributed to effective infection control protocols ^16 17^. Consistent use of appropriate PPE has been shown to reduce the risk of transmission to healthcare workers when caring for an infectious patient ^18^. This may explain why ITU staff in our study (2.7% vs 19.1%, RR 0.1418 95% CI 0.0204 to 0.9869, p 0.0485) had a lower risk of infection, as higher rated PPE was initially recommended for these areas compared to a standard ward. Shortages of masks and inadequate PPE within the NHS during the first wave of COVID-19 were well documented may have contributed to the increased seropositivity of healthcare workers and turned hospitals into reservoirs of potential nosocomial infection^19^.

While transmission of SARS-CoV-2 within households is known to be high, few studies have assessed the particular risk of transmission of SARS-CoV-2 to family members of health care workers ^9^. Lorenzo et al ^20^ found 32.1% of tested family members of healthcare workers were positive for SARS-CoV-2, compared to only 5.3% healthcare workers. They concluded that family members were a greater transmission risk than healthcare professionals. Our study found the opposite, with cohabitees with a positive HCW contact being six times more likely to contract SARS-CoV-2, compared to those without. This suggests that the infection control policies in place at the time were failing to stop transmission of SARS-CoV-2 from the hospital to the wider community.

We were only able to invite cohabitees to attend blood sampling at the end of the national lockdown. As a result of the observed decay in Ig seropositivity seen in the healthcare workers, we have likely underestimated the true rate of SARS-CoV-2 transmission to cohabitees. Using an assumed 2.84 fold decrease in Ig positivity (from an estimated 7% prevalence within the general population during the early phase of the study to the 2.5% rate seen in our “control” cohabitees), we can estimate that the true proportion of positive cohabitees living with a seropositive HCW at the height of the first wave could have been as high 44%.

There was no significant difference in the mean neutralising percentage for participants with symptoms compared to asymptomatic (55.58% vs 51.55%, p =0.405, Independent Samples T Test).

In total, there were 56 seropositive HCWs whose cohabitees donated a serum sample. Of these, nine had a seropositive cohabitee and 47 had one or more seronegative cohabitees. We found no significant differences in the levels of neutralization between those seropositive HCWs who presumably transmitted SARS-CoV-2 to their cohabitee (52.8% neutralization (95% CI 46-143%)) and those whose cohabitees remained seronegative (48.9% (95%CI 32-111%)).

A higher proportion of individuals reported COVID-19 symptoms among the seropositive HCW whose cohabitees seroconverted (55.6 % (95%CI 27-81%)) than those whose cohabitees remained seronegative (37.5% (95%CI 25-52%)) but the difference did not achieve statistical significance.

Based on our study findings, we would recommend that both HCWs and their cohabitees are included in early vaccination programmes when they are available. Universal screening of healthcare workers may also be beneficial. Apart from the cost and manpower implications of this, the current swabs have a false negative rate of between 2 and 33%^21^. While the majority of COVID-19 cases were symptomatic, the majority of people with symptoms were not seropositive. As the majority of participants that reported symptoms were not COVID-19 positive, excluding them from a caring role on symptoms alone would deprive the NHS of staff needlessly during a time of increased resource need.

### Serological response to SARS-CoV-2

Due to the only recent emergence of SARS-CoV-2, our understanding of antibody responses and its clinical implications with regards to possible immunity following repeat exposure is limited. The longitudinal nature of our study allowed for the assessment of the seroconversion event after exposure to SARS-CoV-2. Our study demonstrates reasonable rates of antibody formation despite many of the participants being asymptomatic. Our study demonstrated a decay in rates of IgG, IgA and IgM in positive participants. Rates of IgG, IgA and IgM were similar.

Promisingly, the majority, but not all, of participants who tested positive for SARS-CoV-2 developed measurable neutralising antibodies several weeks after their initial exposure. While the vast majority of positive cohabitees had detectable neutralising antibodies, no cohabitee mounted a neutralising response of over 50% compared to over half of the HCWs. It is probable that the level of neutralising antibodies fell with time, as the cohabitees were tested for neutralising antibodies approximately two months after most of the HCWs. Wajainberg et al found that 90% of patients who were positive at 1:320 mounted a detectable neutralising antibody response for at least 3 to 5 months at identical titres to our study after contracting SARS-CoV-2^22^. We still do not know the required neutralising response to provide longstanding immunity from recurrent SARS-CoV-2 infection. Animal models have suggested protection is gained from SARS-CoV-2 if neutralising antibodies are able to neutralize 50% of the virus at a serum concentration of 1:200 ^23^; further work is required to delineate the response needed to provide immunity in humans.

Peak levels of IgG (spike protein) and white ethnicity may be associated with increasing levels of neutralising antibodies. We were however unable to demonstrate a significant predictor of who goes on to develop a substantial neutralising antibody response of over 50%. We found variation in antibody response between participants, with some participants with a high and sustained antibody titre failing to develop a correspondingly high neutralisation titre. The current global vaccination programmes could provide a further opportunity to assess who develops long-term neutralisation antibodies. While long-term data is lacking, the presence of neutralising antibodies does appear to decline by three months post exposure ^24^. Long term data regarding potential immunity following exposure, including the long term efficacy of the current vaccinations being deployed, remains lacking ^25^. Further work will be urgently needed establish how long this response lasts, and whether this translates to immunity from repeat SARS-CoV-2 infection.

### Risk factors for seropositivity

The evidence defining who is at the greatest risk of contracting and suffering a negative outcome from COVID-19 is rapidly expanding, with both environmental, viral and patient factors being reported^26 27^. The majority of studies have focused on patients and healthcare workers with serious infections requiring hospitalisation, or focused on population level data relying on positive swabs^3^. Few studies have described the incidence of asymptomatic SARS-CoV-2 within healthcare workers and their close contacts using serological data. Other studies have demonstrated risk factors for developing a more severe COVID-19 infection requiring hospitalisation. Identified risk factors for negative outcomes have included black ethnicity, age and male gender^28^. In our study participants presented were largely either asymptomatic or had minimal symptoms. We identified that participants of a black ethnic background were at an increased risk of contracting SARS-CoV-2, independent of social deprivation, gender, age, medical comorbidities and ITU role. This supports the observation that NHS workers of an ethnic minority background are at an increased risk of contracting SARS-CoV-2 ^29^.

### Limitations

There are limitations to this study. A major drawback in terms of assessing the factors affecting transmission is the number of seropositive cohabitees, i.e., putative transmission events. Although we found a considerably larger proportion (55.6%) of symptomatic seropositive HCW has seropositive cohabitees than those who had seronegative cohabitees (37.5%) there are only n=9 pairs where both the HCW and cohabitee were seropositive. This highlights the need for larger HCW-cohabitee pair studies to investigate the risks of transmission. In addition, no single assay has perfect sensitivity and specificity^30^. Few participants had tested positive on nasal SARS-CoV-2 PCR. Routine screening of staff members was not in force during the study period so our results could not be compared to swab or other independently collected results. Our cohort was largely asymptomatic or had low-level symptoms; further study is needed to confirm the generalisability of our results to patients with more severe cases of COVID-19. The cohabitee cohort also only donated a single sample. We were therefore unable to confirm the direction of SARS-CoV-2 transmission as it is possible that the positive cohabitees actually transmitted the virus to the HCW. This raises an additional risk of viral spread, as healthcare workers infected by their cohabitees may transmit the virus into “green” COVID-19 free areas of the hospital. In addition, as the single blood sample was taken later in the study for the cohabitees; based on the observed decay of immunoglobulin levels seen in our study, it is possible that a greater proportion of cohabitees were seropositive earlier during the first wave of the pandemic. We have generated a range of estimates for the likely “true” rate of cohabitee seroprevalence taking into account the observed decay in Ig levels seen in both the healthcare worker cohort and in other studies^31^. Lastly, other immunity pathways may contribute towards immunity to recurrent infection. Further work on the cohort will include assessment of T-lymphocytes and genotyping of both seropositive and seronegative participants.

## Conclusion

Healthcare workers and their cohabitees appear to be an important factor for transmission from hospital to the community. As asymptomatic transmission was common, these groups should be viewed as a priority for regular screening and vaccination.

## Data Availability

Annonymised data is available on request, subject to institutional and ethical approval for release

## Acknowledgements

We would like to acknowledge the contribution of Miss Weronika Rychlik who helped to process the blood samples.

